# Complex Patient Perspectives on Evolving Diverticulitis Treatment

**DOI:** 10.1101/2023.06.26.23291565

**Authors:** Annie Altman-Merino, Kemberlee Bonnet, David Schlundt, Jessie Wrenn, Wesley H. Self, Elisa J. Gordon, Alexander T. Hawkins

## Abstract

**Background:** Despite evidence that antibiotics may not be necessary to treat acute uncomplicated diverticulitis, they remain the mainstay of treatment in the United States. A randomized controlled trial evaluating antibiotic effectiveness could accelerate implementation of an antibiotic-free treatment strategy, but patients may be unwilling to participate.

**Objective:** This study aims to assess patients’ attitudes regarding participation in a randomized trial of antibiotics versus placebo for acute diverticulitis, including willingness to participate.

**Design:** This is a mixed-methods study with qualitative and descriptive methods.

**Settings:** Interviews were conducted in a quaternary care emergency department and surveys were administered virtually through a web-based portal.

**Patients:** Patients with either current or previous acute uncomplicated diverticulitis participated.

**Interventions:** Patients underwent semi-structured interviews or completed a web-based survey.

**Main Outcome measures:** Rates of willingness to participate in a randomized controlled trial was measured. Salient factors related to healthcare decision-making were also identified and analyzed.

**Results:** Thirteen patients completed an interview. Reasons to participate included a desire to help others or contribute to scientific knowledge. Doubts about the efficacy of observation as a treatment method were the main barrier to participation. In a survey of 218 subjects, 62% of respondents reported willingness to participate in a randomized clinical trial. “What my doctor thinks,” followed by “What I’ve experienced in the past” were the most important decision-making factors.

**Limitations:** There is possible selection bias inherent to using a study to evaluate willingness to participate in a study. Also, the population sampled was disproportionately White compared to the population affected by diverticulitis.

**Conclusions:** Patients with acute uncomplicated diverticulitis maintain complex and varying perceptions of the use of antibiotics. Most surveyed patients would be willing to participate in a trial of antibiotics versus placebo. Our findings support a trial’s feasibility and facilitate an informed approach to recruitment and consent.

## Introduction

Diverticular disease is one of the most prevalent conditions in the United States, and its incidence is increasing. Acute diverticulitis is a leading cause of emergency department (ED) visits, accounting for over 360,000 visits per year^1^. Despite evidence that antibiotics are not necessary to treat acute uncomplicated diverticulitis (AUD)^2–4^ and models of diverticulitis pathophysiology suggesting an inflammatory process rather than an infectious one^5, 6^, antibiotics remain the mainstay of treatment in the US.

Unnecessary use of antibiotics is harmful. Antibiotic misuse exacerbates antibiotic resistance, accounting for more dangerous infections and difficult-to-eradicate pathogens. In the US, over 2.8 million antibiotic-resistant infections develop per year, resulting in over 35,000 associated deaths^7^. Overuse of antibiotics leads to longer hospital stays, more readmissions, and ultimately higher mortality rates due to infectious disease^8^. Antimicrobial resistance due to overuse carries a high economic burden of $55 billion per year in the US^7^. Management of uncomplicated diverticulitis without antibiotics represents an opportunity to educate prescribers and reduce antibiotic prescriptions for one of the leading causes of ED visits in the US.

Despite this, antibiotics remain the standard treatment in the US. Three randomized controlled trials in Europe have demonstrated non-inferiority of symptomatic and supportive care without antibiotics compared to treatment with antibiotics for the outcomes of perforation and abscess formation, median hospital stay, and diverticulitis reccurance^2–4^. Guidelines from the American Gastroenterological Association, American Society of Colon and Rectal Surgeons, and American College of Physicians state that uncomplicated cases can be treated without antibiotics^9–12^. The reasons for continued antibiotic use are multifactorial, including lack of physician awareness of updated literature and guidelines, physician fear of medicolegal consequences of “observation,” and patient expectation of treatment with antibiotics^13^. However, the threat of antibiotic resistance is genuine, and continued inappropriate use of antibiotics is antithetical to a rigorous effort to curtail resistance.

A North American randomized controlled trial of antibiotics versus no antibiotics for AUD that demonstrated non-inferiority of an antibiotic-free approach would likely help decrease superfluous antimicrobial use for uncomplicated diverticulitis^13^. This study aims to assess patients’ perceptions and attitudes regarding participation in a randomized controlled trial of antibiotics versus no antibiotics for AUD and evaluate willingness to participate in such a trial.

## Materials and Methods

This mixed-methods study assessed patient perspectives through focused interviews followed by a survey of a separate, larger cohort to quantify themes that emerged from the interviews. Vanderbilt University Medical Center Institutional Review Board approved this study with allowance of verbal consent for the focused interviews (IRB #2210530) and a waiver of informed consent for surveys (IRB #221611).

### Focused Interviews

Semi-structured interviews were conducted both in-person in the Vanderbilt University (VUMC) ED and over the phone. Individuals eligible to participate in in-person interviews included adult (age 18-90 years), English-speaking patients presenting to the VUMC ED with AUD and adequate cognitive capacity to participate. Exclusion criteria for this group included complicated diverticulitis such as bleeding, abscess, or perforation; asymptomatic diverticulitis found incidentally on CT scan; history of colon or rectal cancer, Crohn’s disease, or ulcerative colitis; end-stage renal disease; or previous colectomy. Individuals eligible to participate in phone interviews included adult English-speaking patients with a history of AUD treated at VUMC. Interviews were conducted from October 2022 to December 2022. Once the study team recognized that no novel themes emerged during interviews, it was determined that thematic saturation was reached, and the survey phase concluded^14^.

The interview guide was created with support from the VUMC Qualitative Research Core^15^. It was designed to allow patients to reflect on their expectations, experiences and opinions about diverticulitis using four open-ended questions followed by four focused questions to obtain thoughts about side effects of antibiotics, antibiotic resistance, and treatment guidelines (Full interview guide Supplemental Material 1).

Subjects were identified using natural language processing of radiologists’ interpretations of computed tomography (CT) scans to identify Unified Medical Language System concepts representing diverticulitis. Study personnel were sent an email alert within minutes of radiology interpretation and reviewed the CT scan, radiology interpretation, and patient chart to ensure eligibility. The interviewer (AAM), a female medical student who underwent qualitative interview training with the Qualitative Research Core, approached eligible participants and obtained verbal consent for participation. The interviewer had no prior relationship to patients and disclosed that she is a medical student with interest in the research topic. Interviewees were not compensated. Interviews were recorded using a Sony ICD-PX370 digital voice recorder. Phone interview participants were identified through an electronic medical record query of patients seen in the VUMC Colon and Rectal Surgery Clinic. The interviewer called participants and obtained verbal consent for participation.

### Qualitative Analysis

Audio recordings of interviews were transcribed using an online transcription service (https:\\rev.com) and transcripts were verified manually and not shared with participants. Coding and thematic analysis were conducted by AAM. Interview content was analyzed using open coding and thematic analysis. According to the grounded theory approach, theories were derived from raw data by generating and applying codes^16^. The first five transcripts were reviewed, and each line of text was ascribed one or more codes which were derived inductively from the data. After review of the first five transcripts, codes were distilled into a codebook which was referenced to code the remaining transcripts. Thematic analysis was conducted through four steps of qualitative analysis identified by Green et al - immersion in the data, coding, creating categories, and identification of themes^17^. Qualitative data collection and analysis were performed in accordance with the Standards for Reporting Qualitative Research^18^.

### Survey Methodology

Qualitative analysis of interview data informed the development of a web-based survey to quantify attitudes and beliefs of a separate, larger cohort. Individuals eligible to participate in surveys included patients with a history of diverticulitis. Exclusion criteria were a history of colon and rectal cancer, Crohn’s disease or ulcerative colitis, end-stage renal disease, or previous colectomy.

Using the coded transcripts, potential survey items reflecting patient opinions about antibiotics and treatment of AUD were written. They were organized into an online survey with three sections, each with five items that participants were asked to rank from least important to most important. The survey was reviewed and revised by the research team and beta tested by both physicians and patients. It was distributed as an online survey using REDCap^19^. Additional free text items asked participants to describe their reasons for being willing or unwilling to participate in a randomized clinical trial. (Supplemental Material 2).

Survey participants were identified using social media, VUMC’s electronic health record (EHR), and ResearchMatch, a national health volunteer registry supported by the US National Institutes of Health as part of the Clinical Translational Science Award program. An IRB-approved recruitment message was posted on diverticulitis-related social media pages and sent to eligible ResearchMatch volunteers (Supplemental Materials 3). Eligible participants identified via ICD-10 codes through the EHR were contacted with an IRB-approved message through My Health at Vanderbilt, VUMC’s patient health portal. Participants were given the option to enter a raffle to win a $100 gift certificate upon completion of the survey. Participants were screened for eligibility through a REDCap survey and given access to the study survey if they met eligibility.

### Survey Data Analysis

Descriptive statistics were performed to examine frequencies, means, and standard deviations in survey data. Ranked factors were viewed by number of times ranked “most important” and number of times ranked “least important.” Qualitative analysis was applied to participants’ explanations of willingness or unwillingness to participate with an open coding and thematic analysis approach similar to the methods described in the focused interview section.

## Results

### Participant Characteristics

Thirteen patients completed an interview: nine in the ED with an episode of AUD and four over the phone without a current episode (100% participation for in-person interviews, 67% for phone interviews). All were interviewed alone and only once. Most participants were female (62%, n=8/13) and non-Hispanic White (85%, n=11/13). Detailed patient characteristics are presented in Table 1. Participants had a mean age of 62 years. Mean interview duration was 16 minutes (SD 6 minutes).

**Table 1.**
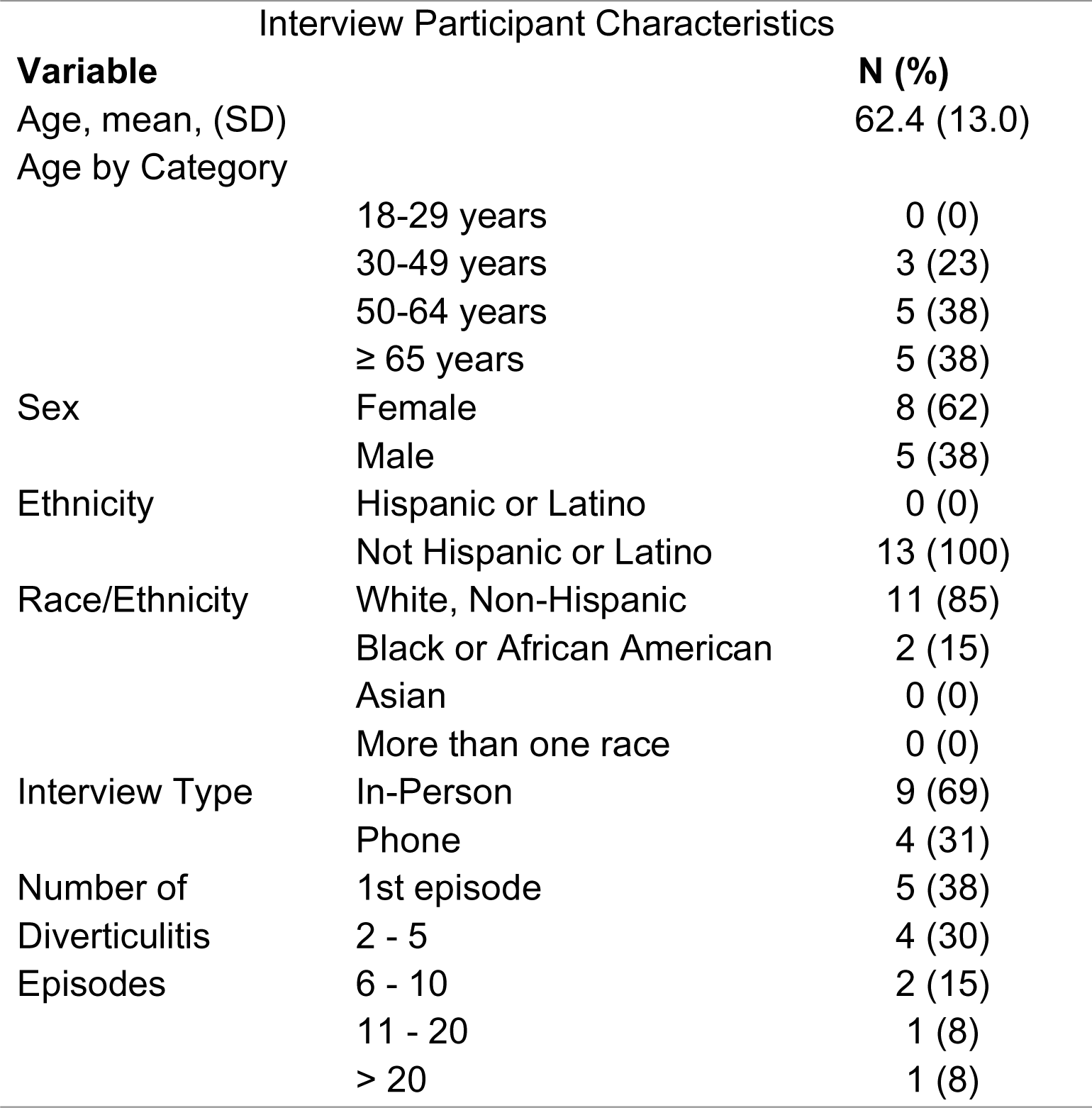
Characteristics of interview participants.

### Themes

Two salient themes that emerged were sources of information used in decision making and factors contributing to reluctance or desire to participate in a trial. Information sources guiding the decision-making process included personal experiences along with provider recommendations. Patients reporting reluctance to participate in a trial commonly expressed doubts about the efficacy of observation as a treatment strategy. Themes regarding willingness to participate were desire to avoid antibiotics in the future, to help others, and to contribute to public knowledge. Figure 1 demonstrates a conceptual model of factors contributing to reluctance or desire to participate. Table 2 includes quotations illustrating each of these themes.

**Figure 1.**
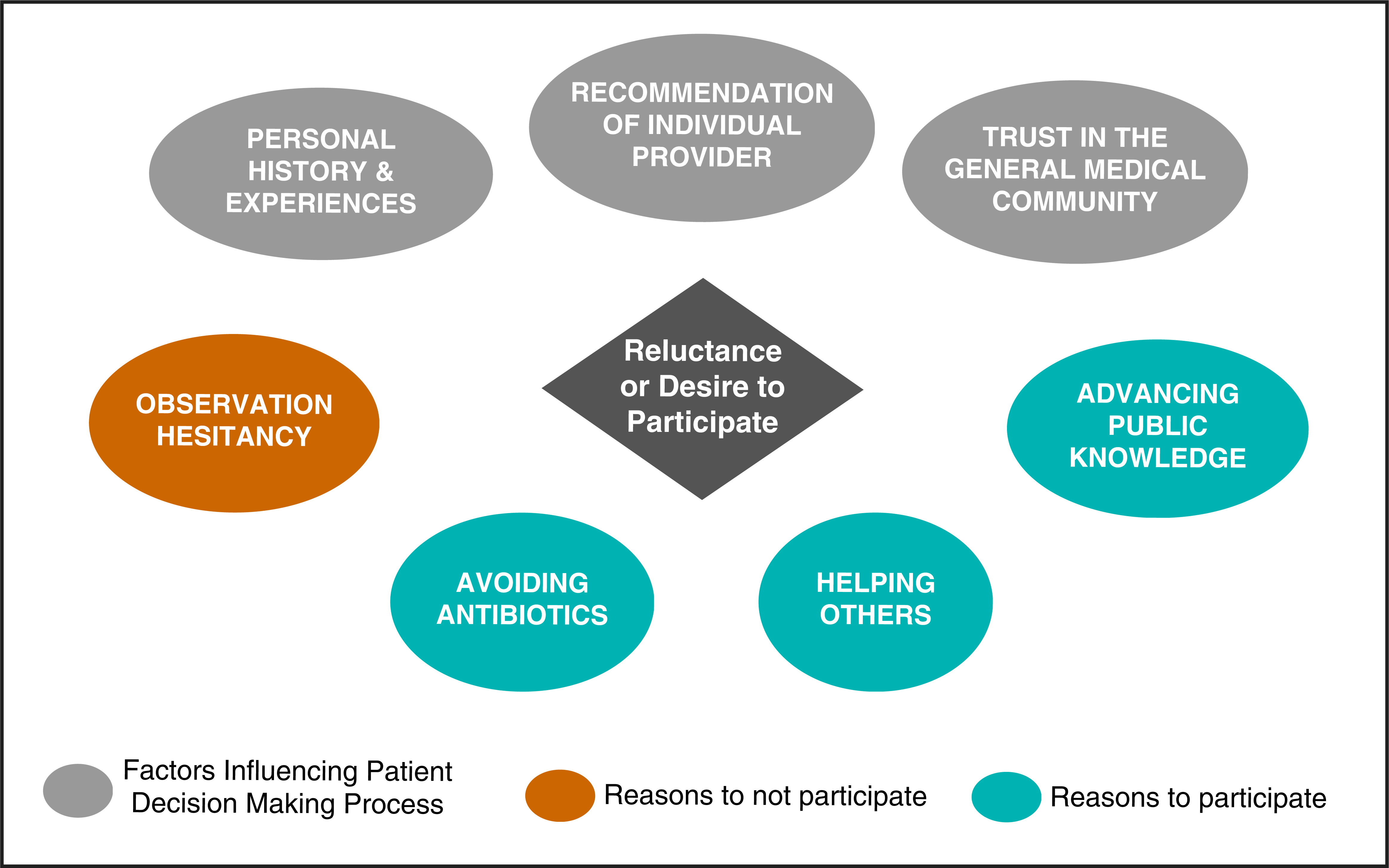
Conceptual model of factors contributing to reluctance or desire to participate.

**Table 2.** Summary of themes and representative quotes.

| Theme | Subtheme | Quotations |
| --- | --- | --- |
| Personal History and Experiences | Antibiotic Effectiveness | With my first episode, I didn't know what it was, and so I had delayed treatment, so it got extremely infected at a bad abscess... And then after that, then I realized that immediate treatment was important. So every time I'd have the symptoms, I would go to the ER as soon as possible...from my experience, catching it as early as possible was critical because delayed treatment clearly, in my experience, made it much worse. (Participant 5) |
|  | Antibiotic Inefficacy | I've had so many episodes and found that [antibiotics] really didn't help. And now my interpretation was that I didn't instantly get better. I got only got better with time as the inflammation settled down. That's my interpretation. (Participant D) |
| Provider Opinion | Known provider | I would rely on my doctor if he told me that this is a situation that can be alleviated through pain management and some other medication or no medication. (Participant 7) |
|  | ED provider | I guess I'm just kind of going on what [the ED doctor] said. If his recommendation was the antibiotic, then I think that would be what I would do. If he said, you can take this or not, and it's going to be the same outcome, I don't know. That might change it if he had said something else. (Participant 6) |
|  | General medical community | I mean, I take it when they tell me I need it and I go blindly...see, as we all do, maybe. (Participant 9) |
| Observation Hesitancy |  | That would be my number one concern is what if it doesn't work? How long do I have to stay in pain before we can go back to what works? (Participant 5) |
| Avoiding antibiotics in the future | Antibiotic hesitancy | I don't ever like to take an antibiotic unless I have to. Okay. Now I'm taking one drug right now that I... I have to take, but I take it, and it has had side effects. So as long as I've been on it, and it's called oh gosh... And when I first got on it, I had such severe diarrhea, I couldn't leave the house about three or four hours. (Participant C) |
|  | Healthcare hesitancy | I used to be involved in the fitness industry and there was a lot of people that were my associates that were very into nutrition, and they believed in healing yourself through healthy eating and exercise and didn't like medication.... So I think we overdo it and I think we damage our immune systems. (Participant 1) |
| Public Impact | Helping Others | I think research is research and as we progress, medicine progresses. So I think whatever needs to be done to, for the betterment of everyone, is what needs to be done. (Participant 8) |
|  | Advancing public knowledge | I mean, on one hand I want to participate in anything it's going to help advance treatment. (Participant 2) |

### Sub-Theme 1: Personal History and Experiences

Participants referenced personal experiences of disease episodes when considering participation in a trial and chronicled a recurring sequence of events: diagnosis of diverticulitis, antibiotic administration, and subsequent symptom resolution. Recognizing and reflecting on the pattern of symptom resolution after antibiotics influenced how participants considered participation in a trial where they may not receive antibiotics.

> *So to be honest, it seems that from my experience, and I know it’s anecdotal, it seems [to] suggest there was a physiological change from the medication. (Participant 2)*

Participant 2 reflected on his past experiences, and by the end of the discussion, had drawn upon them to reach a decision:

> *Well, talking it out, it’s kind of helped me process and see that from a very first episode, it seems like the amoxicillin was effective… So actually, just talking to you, I started to lean more towards staying with amoxicillin. (Participant 2)*

Patients’ perceptions of antibiotic efficacy in the past, whether perceived as high or low efficacy, informed decisions about participation in a trial. This individual perceived that antibiotics were effective during previous episodes, which precipitated reluctance to participate in a trial. Given the recurring and episodic nature of diverticular disease, personal experience is particularly salient to decisions about disease management.

### Sub-Theme 2: Influence of Providers on Participant Perceptions

Participants considered providers’ recommendations about whether antibiotics would be beneficial as central to their perceptions of diverticulitis treatment. Some participants identified their individual primary care provider or the ED provider as trusted sources of guidance about treatment options.

> *I think I would need to talk to my doctor, my family doctor. I think that would be my best bet. (Participant A)*

Others invoked the authority of providers as a collective medical community who have accepted antibiotics as the standard treatment.

> *You just trust the science or whatever … it doesn’t matter which doctor, everybody prescribes the antibiotic regardless …. So I’ve never questioned it because that’s what they say it takes. (Participant 5)*

### Sub-Theme 3: Observation Hesitancy

Most participants who were unwilling to participate in a trial doubted the efficacy of observation as a treatment strategy, believing that antibiotics are necessary for recovery. These participants tended to view the omission of antibiotics as the omission of treatment.

> *At least when I have it, I mean, I can’t imagine if there was no treatment, because I’ve had to wait before to get treatment and it didn’t get any better. (Participant 6)*

The interviewer informed participants that the placebo arm would receive treatment in the form of supportive care, pain control, and close observation. However, some participants interpreted the lack of antibiotics as “no treatment.”

> *So if I’m getting the sugar pill then I’m in the control group, does that mean I’m not getting any treatment at all? (Participant B)*

### Sub-Theme 4: Desire to Avoid Antibiotics in the Future

Aversion towards antibiotics contributed to both reluctance and desire to participate. Some patients feared placement in the antibiotic study group and others saw participation in the study as a means of avoiding antibiotics in the future.

> *But I never take an antibiotic unless I have to. I just am afraid it would - you take too many and it won’t work. (Participant C)*

Concern about taking any medication and aversion to unnecessary medical treatment in general were commonly shared sentiments.

> *Well, I think that people get over-medicated sometimes, I do. (Participant 4)*

### Sub-Theme 5: Helping Others and Advancing Public Knowledge

The public impact of study participation, including advancing public knowledge and helping others, were primary drivers of patients’ willingness to participate.

> *I struggle with this for 10 years and any advancement in the treatment, I’d be glad to be part of that. Yeah, it would help a lot of people, I would think. (Participant 2)*

### Survey Quantitative Results

Two hundred and eighteen participants completed the survey. Participation rate by method is demonstrated in Figure 2. Mean participant age was 57.9 years (SD 13.3 years), and most were female (61.9%), and non-Hispanic White (69.7%). Detailed patient characteristics are presented in Table 3.

**Figure 2.**
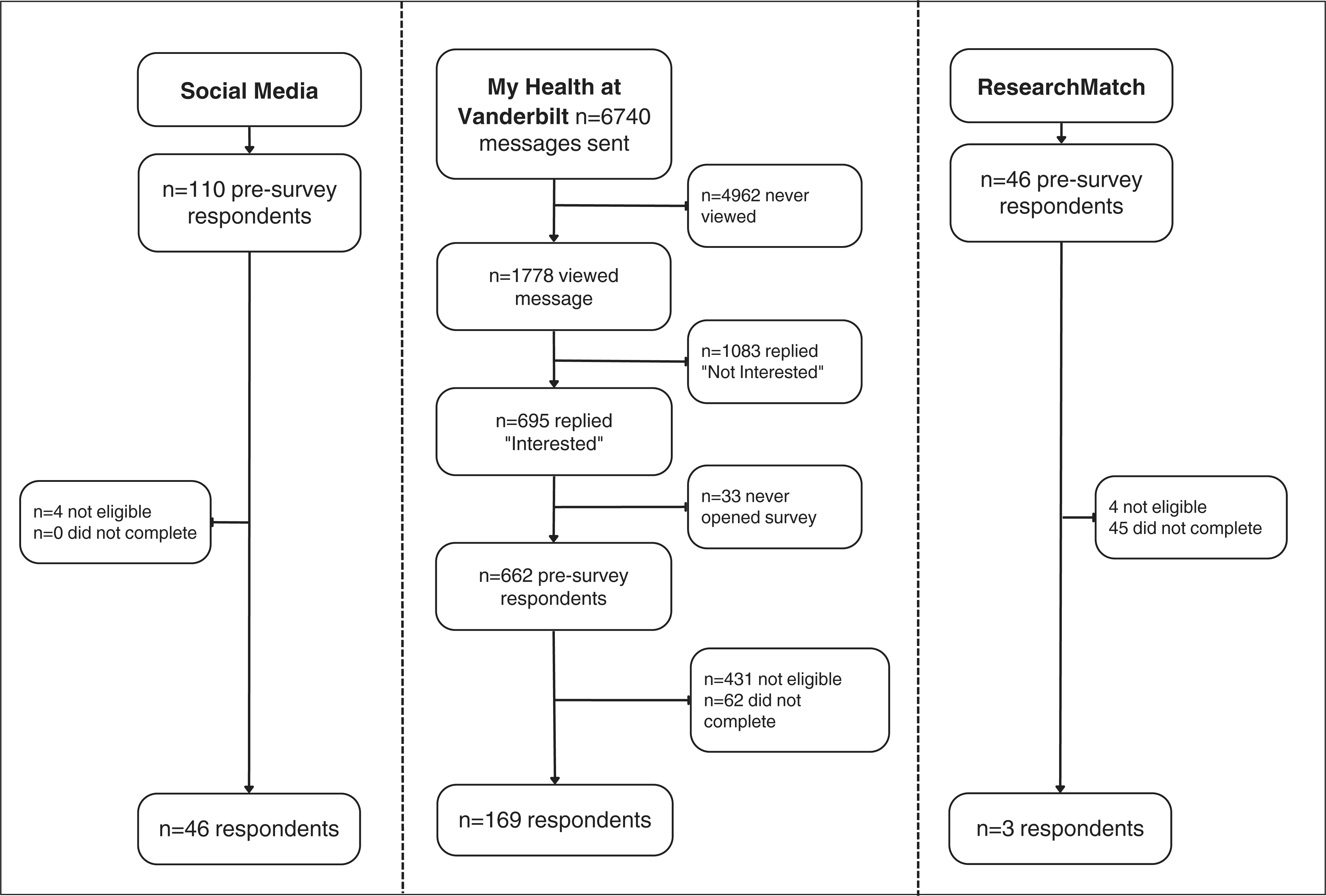
Survey participation rate by recruitment method.

**Table 3.** Characteristics of survey participants.

Survey Participant Characteristics
| **Variable** |  | **N (%)** |
| --- | --- | --- |
| Age, mean, (SD) |  | 57.9 (13.3) |
| Age by Category | 18-29 years | 3 (1.4) |
|  | 30-49 years | 55 (25.2) |
|  | 50-64 years | 68 (31.2) |
|  | ≥ 65 years | 81 (37.2) |
| Sex | Female | 135 (61.9) |
|  | Male | 77 (35.3) |
|  | Other/Prefer not to say | 2 (0.9) |
|  | Not reported | 4 (1.8) |
| Ethnicity | Hispanic or Latino | 12 (5.5) |
|  | Not Hispanic or Latino | 191 (87.6) |
|  | Unknown | 3 (1.4) |
|  | Not reported | 12 (5.5) |
| Race/Ethnicity | White, Non-Hispanic | 152 (69.7) |
|  | Black or African American | 9 (4.1) |
|  | Asian | 1 (0.5) |
|  | More than one race | 4 (1.8) |
|  | Not reported | 31 (14.2) |
| Recruitment Site | ResearchMatch | 21 (14) |
|  | MHAV | 146 (67) |
|  | Facebook | 4 (3) |
|  | Reddit | 41 (27) |
| Number of Diverticulitis Episodes | 1 - 5 | 172 (78.9) |
|  | 6 - 10 | 32 (14.7) |
|  | 11 - 20 | 10 (4.6) |
|  | > 20 | 2 (0.9) |

One Hundred and thirty-five respondents (62%) reported willingness to participate in a trial of antibiotics versus placebo during an episode of AUD. Figure 3 illustrates the number of times each factor was ranked most important. The two leading reasons to participate included “Helping make guidelines for treatment better” (48%) and “Avoiding antibiotics” (22%).

**Figure 3.**
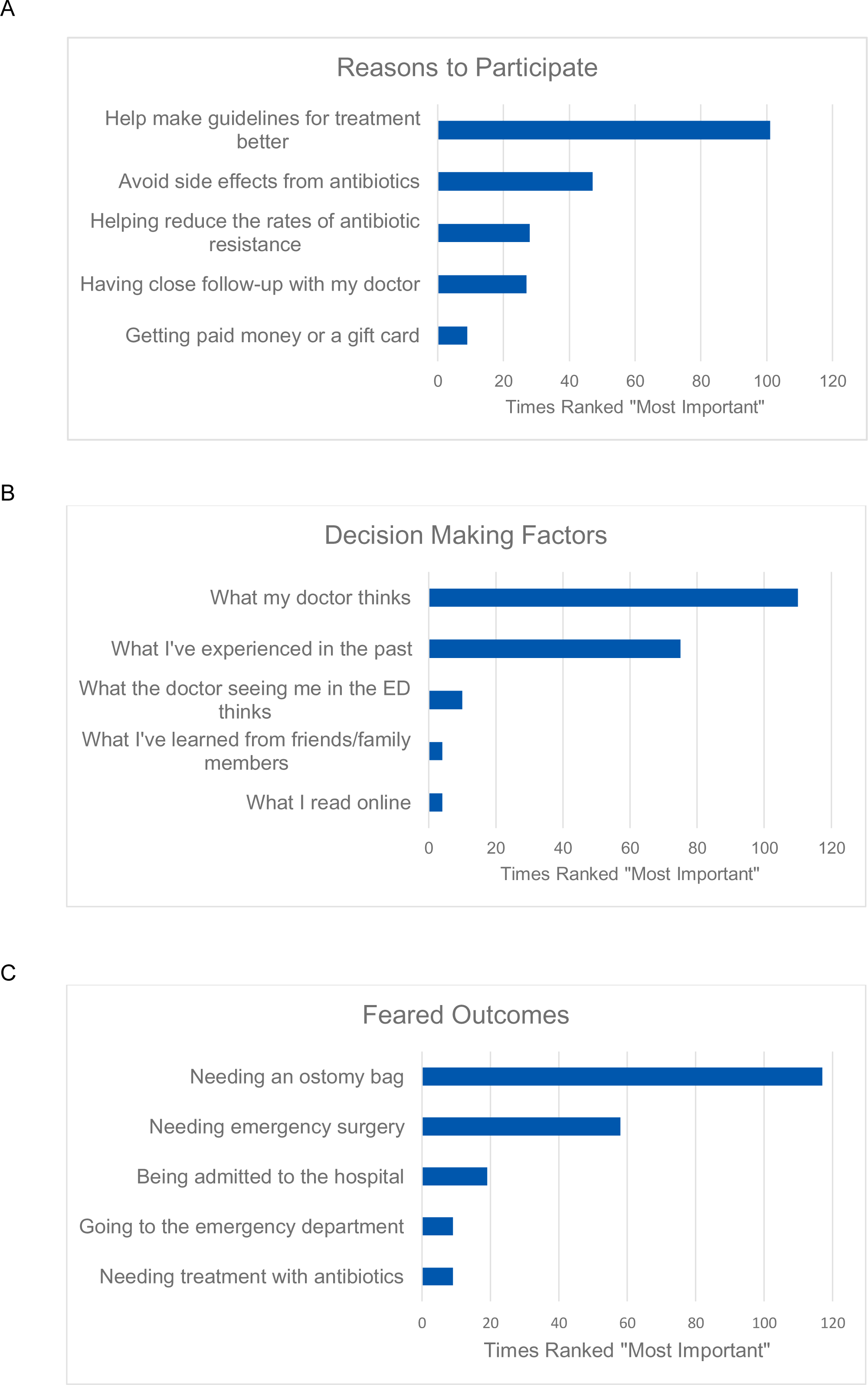
Survey factors represented by times ranked “Most Important,” including A, reasons to participate in a randomized controlled trial of antibiotics versus placebo, B, factors used to make healthcare decisions, and C, feared outcomes of diverticulitis.

When prompted to rank the importance of various factors used to make healthcare decisions, the leading factors were “What my doctor thinks,” (56%) and “What I’ve experienced in the past” (36%). Only 5% reported “What the doctor seeing me in the Emergency Department thinks” as most important. “Needing an ostomy bag” was the most feared outcome of diverticulitis (55%).

### Qualitative Results of Surveys

Among 83 who reported unwillingness to participate, 67 explained their response. These explanations largely fall into two categories: fear of an antibiotic-free treatment and aversion to antibiotics themselves.

> *I have had diverticulitis three times and each time it was cleared up with antibiotics. The thought of getting a placebo scares me. (Male in their 60s)*

Fear of not receiving antibiotics contrasts with preference for a non-antibiotic strategy.

> *Don’t trust antibiotics. Makes it worse. Longer to heal. (Male in their 60s)*

Those who were willing to participate specified a desire to help others, contribute to diverticulitis-related research, or understand the disease themselves.

> *I like to help and would like a better understanding of diverticulitis. (Female in their 40s)*

## Discussion and Conclusions

. Considering that patient hesitancy to participate in a trial of antibiotics versus placebo has been a major barrier to its execution, understanding patients’ opinions is essential to conducting a trial and ultimately implementing a change in the standard treatment of diverticulitis. Both qualitative literature and literature regarding patient perspectives are increasingly recognized as important for informing treatment strategies and understanding trial recruitment^14, 20–22^. This mixed-methods study assessed patients’ perceptions of diverticulitis treatment options through semi-structured interviews with patients in addition to a survey of a larger cohort, and found that most patients with a history of diverticulitis report willingness to participate in a trial of antibiotics versus placebo for AUD. It also identified factors that patients consider when making decisions related to their healthcare, with their doctor’s recommendation being the most important.

Over 60% of survey respondents reported willingness to participate in a trial of antibiotics versus placebo for AUD. This enthusiasm for trial participation would support robust enrollment in an actual trial and scientific integrity. Our findings support the feasibility of such a trial and can be applied during its planning and execution to facilitate patient-centeredness in methods and analysis. This number is especially encouraging when compared to participation rates in similar US surgical trials that have recruited from the ED. For example, the CODA trial, comparing antibiotics with appendectomy for appendicitis, had a 31% enrollment rate among eligible patients^23^.

Confidence in providers was evident in conversations with patients and survey responses. ED provider opinions carried weight particularly in interviews, where these providers were the first to communicate diagnoses and present treatment options. Interviewees also expressed confidence in doctors and the medical community to make the best decision for them. These findings highlight the potential for providers to affect patient perspectives on the use of an antibiotic-free strategy.

A key strength of this study is that interviews simulated enrollment in an RCT: most patients were recruited in the ED at the time of diagnosis, before either discharge or hospital admission. This recruitment approach provided a unique view into patient perceptions of treatment options at the time that they would be enrolled, and therefore reflected authentic concerns, doubts, and questions that typically arise in that context.

Our study has limitations. There is the possibility of selection bias, which is inherent to conducting a study to evaluate willingness to participate in a study. However, all individuals approached in-person in the ED agreed to participate in an interview, indicating that the population was not skewed towards those who are more inclined to participate. The population sampled was disproportionately White compared to the population affected by diverticulitis, meaning that the perspectives of some voices underrepresented in biomedical research are left out. Another limitation was that survey respondents did not have the opportunity to engage in dynamic discussion with the research team. Some participants misunderstood the explanation of the trial and indicated unwillingness to undergo randomization because they were not currently experiencing symptoms. Consequently, participants’ willingness may have been underestimated.

In conclusion, the most important factors guiding patients’ decision making about diverticulitis treatment include recommendations by trusted providers and perceptions of observation as ineffective treatment. These factors may be modulated over time as providers accept an antibiotic-free approach and affirm its legitimacy to patients. A US-based randomized controlled trial of antibiotics versus placebo for acute uncomplicated diverticulitis is feasible in terms of patient recruitment, and even anticipated by patients.

## Data Availability

All data produced in the present study are either contained in the manuscript or available upon reasonable request to the authors.

## Author contribution

AAM designed and executed all study procedures including manuscript drafting and editing. KB assisted interview guide design, qualitative training of primary author, and manuscript writing. DS supported qualitative analysis including writing of the corresponding manuscript section. JW and WHS assisted with interviewee identification through the emergency department and contributed to writing and editing. EG supported qualitative analysis and contributed to writing and editing. ATH oversaw all study procedures and contributed to writing and editing. All authors reviewed the final manuscript.

## Acknowledgements

We thank Dario Giuse, Dr. Ing, MS, FACMI, for his contribution to the study.

## References

1. Bollom A, Austrie J, Hirsch W, et al. Emergency department burden of diverticulitis in the USA, 2006-2013. Dig Dis Sci. 2017;62:2694–2703.

2. Chabok A, Påhlman L, Hjern F, Haapaniemi S, Smedh K; AVOD Study Group. Randomized clinical trial of antibiotics in acute uncomplicated diverticulitis. Br J Surg. 2012;99(4):532–539. doi:10.1002/bjs.8688

3. Daniels L, Ünlü Ç, de Korte N, et al. Randomized clinical trial of observational versus antibiotic treatment for a first episode of CT-proven uncomplicated acute diverticulitis. Br J Surg. 2017;104(1):52–61. doi:10.1002/bjs.10309

4. Mora-López L, Ruiz-Edo N, Estrada-Ferrer O, et al. Efficacy and Safety of Nonantibiotic Outpatient Treatment in Mild Acute Diverticulitis (DINAMO-study): A Multicentre, Randomised, Open-label, Noninferiority Trial. Ann Surg. 2021;274(5):e435–e442. doi:10.1097/SLA.0000000000005031

5. Piscopo N, Ellul P. Diverticular Disease: A Review on Pathophysiology and Recent Evidence. Ulster Med J. 2020;89(2):83–88.

6. Zullo A. Medical hypothesis: speculating on the pathogenesis of acute diverticulitis. Ann Gastroenterol. 2018;31(6):747–749.

7. Centers for Disease Control and Prevention. Antibiotic Resistance Threats in the United States. https://www.cdc.gov/drugresistance/pdf/threats-report/2019-ar-threats-report-508.pdf. 2019.

8. Dadgostar P. Antimicrobial Resistance: Implications and Costs. Infect Drug Resist. 2019;12:3903–3910. Published 2019 Dec 20. doi:10.2147/IDR.S234610

9. Peery AF, Shaukat A, Strate LL. AGA Clinical Practice Update on Medical Management of Colonic Diverticulitis: Expert Review. Gastroenterology. 2021;160(3):906–911.e1. doi:10.1053/j.gastro.2020.09.059

10. Stollman N, Smalley W, Hirano I; AGA Institute Clinical Guidelines Committee. American Gastroenterological Association Institute Guideline on the Management of Acute Diverticulitis. Gastroenterology. 2015;149(7):1944–1949. doi:10.1053/j.gastro.2015.10.003

11. Hall J, Hardiman K, Lee S, et al. The American Society of Colon and Rectal Surgeons Clinical Practice Guidelines for the Treatment of Left-Sided Colonic Diverticulitis. Dis Colon Rectum. 2020;63(6):728–747. doi:10.1097/DCR.0000000000001679

12. Qaseem A, Etxeandia-Ikobaltzeta I, Lin JS, et al. Diagnosis and Management of Acute Left-Sided Colonic Diverticulitis: A Clinical Guideline From the American College of Physicians. Ann Intern Med. 2022;175(3):399–415. doi:10.7326/M21-2710

13. Garfinkle R, Sabboobeh S, Demian M, Barkun A, Boutros M; Management of Uncomplicated Diverticulitis (MUD) Collaborative. Patient and Physician Preferences for Antibiotics in Acute Uncomplicated Diverticulitis: A Delphi Consensus Process to Generate Noninferiority Margins. Dis Colon Rectum. 2021;64(1):119–127. doi:10.1097/DCR.0000000000001815

14. Saunders B, Sim J, Kingstone T, et al. Saturation in qualitative research: exploring its conceptualization and operationalization. Qual Quant. 2018;52(4):1893–1907. doi:10.1007/s11135-017-0574-8

15. Vanderbilt University Medical Center (2023). Vanderbilt University Qualitative Research Core. Vanderbilt Institute for Medicine and Public Health. Retrieved May 3, 2023, from https://www.vumc.org/hsr/qualitative-research-core

16. Chun Tie Y, Birks M, Francis K. Grounded theory research: A design framework for novice researchers. SAGE Open Med. 2019;7:2050312118822927. Published 2019 Jan 2. doi:10.1177/2050312118822927

17. Green J, Willis K, Hughes E, et al. Generating best evidence from qualitative research: the role of data analysis. Aust N Z J Public Health. 2007;31(6):545–550. doi:10.1111/j.1753-6405.2007.00141.x

18. O’Brien BC, Harris IB, Beckman TJ, Reed DA, Cook DA. Standards for reporting qualitative research: a synthesis of recommendations. Acad Med. 2014;89(9):1245–1251. doi:10.1097/ACM.0000000000000388

19. Harris PA, Taylor R, Thielke R, Payne J, Gonzalez N, Conde JG. Research electronic data capture (REDCap)--a metadata-driven methodology and workflow process for providing translational research informatics support. J Biomed Inform. 2009;42(2):377–381. doi:10.1016/j.jbi.2008.08.010

20. Ndjaboue R, Chipenda Dansokho S, Boudreault B, et al. Patients’ perspectives on how to improve diabetes care and self-management: qualitative study. BMJ Open. 2020;10(4):e032762. Published 2020 Apr 29. doi:10.1136/bmjopen-2019-032762

21. Moorcraft SY, Marriott C, Peckitt C, et al. Patients’ willingness to participate in clinical trials and their views on aspects of cancer research: results of a prospective patient survey. Trials. 2016;17:17. Published 2016 Jan 9. doi:10.1186/s13063-015-1105-3

22. Ferrell B, Williams AC, Borneman T, Chung V, Smith TJ. Clinical Trials: Understanding Patient Perspectives and Beliefs About Treatment. Clin J Oncol Nurs. 2019;23(6):592–598. doi:10.1188/19.CJON.592-598

23. CODA Collaborative, Flum DR, Davidson GH, et al. A Randomized Trial Comparing Antibiotics with Appendectomy for Appendicitis. N Engl J Med. 2020;383(20):1907–1919. doi:10.1056/NEJMoa2014320

