## Supplemental Material 1 for "Complex Patient Perspectives on Evolving Diverticulitis Treatment"

Supplemental Material 1. Full interview guide.

For a long time, doctors have treated episodes of acute diverticulitis, like you’re having now, with antibiotics. But some studies have shown that doctors might not need antibiotics to treat it. We are planning to do a study to compare treatment with antibiotics to treatment without antibiotics, just medicine for pain. In studies like this, patients are randomly assigned to get either antibiotics or a “placebo” (“sugar pill”).   They do not know which one they are getting until after the study ends. Before we do this, we’d like to learn more about the experiences of people with diverticulitis and what they think about this. I’d like to ask you a few questions about your experiences and what’s most important to you. Is that ok?

 Is it okay with you if I record the interview? Your name **will not** be associated with what we record or anything that you say.

1. How would you feel about being randomly assigned to take either antibiotics or no antibiotics for treatment of your diverticulitis?
   a. Why or why not?
   b. What would be your concerns about participating?
2. Can you tell me about your experience with diverticulitis and how it affects your daily life?
   a. When was your first episode?
   b. How long have you had diverticulitis?
   c. How often do you get symptoms?
3. What symptoms are most bothersome to you?
4. One thing that many researchers who study treatments for diverticulitis have looked at is “time to recovery,” or the time it takes to feel better after an episode. What does the idea of recovery mean to you?
5. Have you heard of antibiotic resistance? If so, can you tell me what you understand?
   a. If “no”: In an infection, there’s a group of bacteria. Antibiotics work by killing bacteria. When we use antibiotics, the weak bacteria die but some stronger ones survive. When doctors give antibiotics too much, the whole group of bacteria gets stronger, and antibiotics don’t work as well. It’s important for doctors to only give you antibiotics when they are necessary. Have you heard about this before? What are your thoughts about taking antibiotics even though using them too much can make them not work as well?
6. Like all medicines, antibiotics have side effects. What are your concerns about the effects of using antibiotics?
7. Guidelines are recommendations by experts based on research and experience with patients that help doctors make decisions. Treatment guidelines now say that in most cases, diverticulitis can be treated without antibiotics. Does this change how you feel about trying treatment without antibiotics?
8. After talking to me, how have your ideas about being randomly assigned to antibiotics or no antibiotics changed?
9. Would you be willing to be randomly assigned to take either antibiotics or no antibiotics for treatment of your diverticulitis as part of a study? (as a reminder, we are not recruiting patients for this now, we just want to know what people think about it).
   a. What would be your concerns about participating?
   b. If the study involved more follow-up visits than usual with your doctor, would this make you more enthusiastic about participating?
   c. If the study involved compensation in the form of money or a gift card, would this make you more enthusiastic about participating?
      i. How much money do you think would be fair?
10. What else would you need to know to decide about whether to participate?

*** Society of American GI and Endoscopic Surgeons, European Association of Endoscopic Surgery, American Gastroenterological Association, ASCRS*
