## Supplemental Material 2 for "Complex Patient Perspectives on Evolving Diverticulitis Treatment"

### Diverticulitis Eligibility Survey

**Thanks for your interest in our study! Please answer the questions below to confirm that you are eligible to participate. If you provide an answer that indicates you are not eligible, a message will appear that the survey has ended.**

1) Have you ever been diagnosed with diverticulitis?
   ◯ Yes
   ◯ No

2) Have you ever been diagnosed with colon cancer or rectal cancer?
   ◯ Yes
   ◯ No

3) Have you ever been diagnosed with Inflammatory Bowel Disease, including Crohn's Disease or Ulcerative Colitis?
   ◯ Yes
   ◯ No

4) Have you ever had a surgery to remove part or all of your colon or rectum?
   ◯ Yes
   ◯ No

5) I certify that I am at least 18 years old and that I give my consent to participate in this study.
   ◯ I consent
   ◯ I do not consent

### Participant Demographic Info

Please complete the questions below to tell us more about yourself.

Thank you!

---

Age

______________________

---

Ethnicity

◯ Hispanic or Latino ◯ NOT Hispanic or Latino ◯ Unknown / Not Reported

---

Race

◯ American Indian/Alaska Native
◯ Asian
◯ Native Hawaiian or Other Pacific Islander
◯ Black or African American
◯ White
◯ More Than One Race
◯ Unknown / Not Reported

---

Gender

◯ Female
◯ Male
◯ Other
◯ Prefer not to say

---

Where did you hear about our study?

◯ ResearchMatch
◯ MyHealth at Vanderbilt message
◯ Facebook
◯ Twitter
◯ Reddit
◯ Other

---

Have you ever been hospitalized for diverticulitis?

◯ Yes
◯ No

---

If yes, about how many times have you been hospitalized for diverticulitis?

______________________

---

About how many episodes of diverticulitis have you had?

______________________

---

About how long ago were you diagnosed with diverticulitis?

◯ Less than 1 year ago
◯ 1-5 years ago
◯ 5-10 years ago
◯ 10-20 years ago
◯ Over 20 years ago

---

Does an immediate family member (parent, sibling, aunt/uncle, cousin) suffer from diverticulitis? If so, please list their relationship to you.

______________________

### Diverticulitis Questions

Please complete the survey below about your experience with diverticulitis.

Thank you!

**We are planning a study to see whether antibiotics are necessary to treat diverticulitis. In this study, people who came to the emergency department with an episode or diverticulitis flare would either take antibiotics or a placebo ("sugar pill") without knowing which one they are taking. These are both safe options and people would only be in the study if their doctors agreed that it was safe for them. Before we do this study, we want to hear from patients with diverticulitis. We want to hear about your experiences and thoughts about participating in a study like this. Completing this survey does not require you to participate in the later study.**

Would you be willing to participate in a study where you are assigned to take either antibiotics or a placebo?

◯ Yes
◯ No

Please explain your answer (optional):

______________________________

On a scale of 1-10, how confident are you that antibiotics are needed to treat diverticulitis? (1 means you do NOT think they are necessary, and 10 means you are certain that they are necessary)

1 ________________________________ 10

*(Place a mark on the scale above)*

**Below are reasons that someone might want to participate. Please rank them in order of most to least important to you. Rank the most important one #1 and the least important one #5.**

| | 1- most important | 2 | 3 | 4 | 5 - least important |
|---|---|---|---|---|---|
| Getting paid money or a gift card | ◯ | ◯ | ◯ | ◯ | ◯ |
| Avoiding side effects* from antibiotics | ◯ | ◯ | ◯ | ◯ | ◯ |
| Having close follow-up with my doctor | ◯ | ◯ | ◯ | ◯ | ◯ |
| Helping reduce the rates of antibiotic resistance** for the good of the public | ◯ | ◯ | ◯ | ◯ | ◯ |
| Helping make guidelines*** for treatment better | ◯ | ◯ | ◯ | ◯ | ◯ |

* The most common side effects of antibiotics used to treat diverticulitis are nausea, vomiting, diarrhea, abdominal pain, and rash. Rare but more serious side effects include C. diff infection, confusion, and seizures.

** When doctors give antibiotics too much, they don't work as well. This is called "antibiotic resistance." It's important for doctors to only give you antibiotics when they are necessary.

*** Guidelines are recommendations by experts based on experiences with patients and research. Doctors use guidelines to help them make decisions about treatment.

What might increase your willingness to participate in a study like this? ______________________

**Please rank the following in order of what is most to least important to you for making decisions about your healthcare. Rank the most important one #1 and the least important one #5:**

| | 1 - most important | 2 | 3 | 4 | 5 - least important |
|---|---|---|---|---|---|
| What my doctor thinks | ○ | ○ | ○ | ○ | ○ |
| What the doctor seeing me in the Emergency Department thinks | ○ | ○ | ○ | ○ | ○ |
| What I read online | ○ | ○ | ○ | ○ | ○ |
| What I've experienced in the past | ○ | ○ | ○ | ○ | ○ |
| What I've learned from friends/family members | ○ | ○ | ○ | ○ | ○ |

**Below are factors you might be concerned about or afraid of when you have diverticulitis. Please rank them in order, with the most concerning/fearsome one #1 and the least concerning/fearsome one #5:**

| | 1 - most concerning | 2 | 3 | 4 | 5 - least concerning |
|---|---|---|---|---|---|
| Being admitted to the hospital | ○ | ○ | ○ | ○ | ○ |
| Needing treatment with antibiotics | ○ | ○ | ○ | ○ | ○ |
| Needing an ostomy bag | ○ | ○ | ○ | ○ | ○ |
| Needing emergency surgery | ○ | ○ | ○ | ○ | ○ |
| Going to the Emergency Department | ○ | ○ | ○ | ○ | ○ |

What else are you concerned about or afraid of when you have diverticulitis? ______________________

### Participant Updater Module.

| | |
|---|---|
| MRN | __________________ |
| Study Status | __________________ |
| First Name | __________________ |
| Last Name | __________________ |
| DOB | __________________ |
| Email | __________________ |
