## Supplemental Material 3 for "Complex Patient Perspectives on Evolving Diverticulitis Treatment"

**Supplemental Material 3. IRB-approved recruitment messages.**

Social Media Recruitment Message

Do you have diverticulitis? Researchers at Vanderbilt University Medical Center are looking for volunteers for a study about patient perspectives on diverticulitis. If you complete the survey, you can enter a raffle to win a $100 gift certificate!

The purpose of this study is to understand patient experiences with diverticulitis and preferences about treating it. We want to hear about your experiences to improve care for patients with diverticulitis. The study involves a 5-minute survey that can be completed online or over the phone.

Survey link: https://redcap.vanderbilt.edu/surveys/?s=WK4KEWE9KT3PDAPP

If you are interested in participating but have questions or concerns or would like to speak to a study team member, you can reach out to me via email at or call me at 615-669-7086.

Research Match Recruitment Message

Do you have diverticulitis? Researchers at Vanderbilt University Medical Center are looking for volunteers for a study about patient perspectives on diverticulitis.

The purpose of this study is to understand patient experiences with diverticulitis and preferences about treating it. We want to hear about your experiences to improve care for patients with diverticulitis. The study involves a 5-minute survey that can be completed online or over the phone.

You may be eligible for this study if you:

- -  Have diverticulitis
- -  Have never been diagnosed with colorectal cancer
- -  Have never been diagnosed with Crohn’s Disease or Ulcerative Colitis
- -  Have never had surgery to remove all or part of your colon

If you select “yes,” you will receive an email from me with more information and a link to the survey.
